# COVID-19 vaccine hesitancy among individuals with cancer, autoimmune diseases, and other serious comorbid conditions

**DOI:** 10.1101/2021.04.06.21254014

**Authors:** Richard Tsai, John Hervey, Kathleen D Hoffman, Jessica Wood, John Novack, Jennifer Johnson, Dana C. Deighton, Brian Loew, Stuart L Goldberg

## Abstract

**Background:** Individuals with comorbid conditions have been disproportionately affected by COVID-19. Since regulatory clinical trials with COVID-19 vaccines excluded those with immunocompromising conditions, few patients with cancer and autoimmune diseases were enrolled. With limited vaccine safety data available, vulnerable populations may have conflicted vaccine attitudes.

**Methods:** To assess the incidence and reasons for COVID-19 vaccine hesitancy and to assess early vaccine safety, we conducted a cross-sectional online survey, fielded January 15, 2021 through February 22, 2021, with international participation (74% USA). A random sample of members of Inspire, an⍰online health⍰community⍰of over 2.2⍰million individuals⍰with comorbid conditions, completed a 55-item online survey.

**Results:** 21,943 individuals completed the survey (100% with comorbidities including 27% cancer, 23% autoimmune diseases, 38% chronic lung diseases). 10% declared they would not, 4% stated they probably would not, and 5% were not sure they would agree to vaccination (hesitancy rate 19%). Factors associated with hesitancy included younger age, female gender, black-Pacific-Island-Native American heritage, less formal education, conservative political tendencies, resistance to masks or routine influenza vaccinations, and distrust of media coverage. 5501 (25%) had received at least one COVID-19 vaccine injection, including 29% of US participants. Following the first injection, 69% self-reported local and 40% systemic reactions, which increased following the second injection to 76% and 67%, respectively, with patterns mimicking clinical trials.

**Conclusion:** Nearly one in five individuals with serious comorbid conditions harbor COVID-19 hesitancy. Early safety experiences among those who have been vaccinated should be reassuring.

**Highlights:**

- Individuals with serious comorbid conditions, including cancer, have been disproportionately affected by COVID-19 and therefore have been prioritized for vaccination
- An online survey of nearly 22,000 individuals with comorbid conditions revealed that nearly one in 5 expressed vaccine hesitancy.
- Reasons for hesitancy in this comorbid population mimicked surveys of the general population.
- Self-reported safety profiles among individuals with comorbid conditions were acceptable, and generally milder than reports in clinical trials among the general population.

## 1. Introduction

The rapid development of safe and effective vaccines against SARS-CoV-2 may stem the global COVID-19 pandemic. However, vaccine hesitancy – the reluctance or refusal to vaccinate – has emerged as a major worldwide public health concern, especially as it may impair the ability to reach herd immunity status. (1-3) An IPOS poll for the World Economic Forum conducted in January 2021 of 15 countries reported vaccine acceptance rates ranging from 86% in Brazil to only 46% in Russia, with the United States ranking 12^th^ (63% vaccine acceptance rate). (4) Over time vaccine acceptance has increased. Serial tracking polls by the Kaiser Family Foundation conducted in the United States report that 55% of adults have either received or will receive as soon as possible the vaccine as of February 2021, up from 47% in mid-January and 34% in early December 2020. However, anti-vaccination sentiment has remained constant over this timeframe with 15% stating that they will definitely not get vaccinated and 7% agreeing only if required. (5) Several studies have explored reasons for COVID-19 vaccine hesitancy, with vaccine-specific concerns (side-effects and efficacy), a need for more information, racial/ethnic biases, political views, general anti-vaccine attitudes or beliefs, and a lack of trust being most commonly cited. (5-9)

Individuals with comorbid conditions have been disproportionately affected by COVID-19. A review of nearly a half-million commercially insured COVID-19 patients noted that although only 51.7% had pre-existing conditions, 83.3% of COVID-19 related deaths occurred among those with comorbidities. The risk of dying from COVID-19 was strongly correlated with the number of comorbidities, nearly doubling with a single condition and increasing 8-fold with five or more conditions. (10) Persons with developmental disorders, congenital and acquired neurologic disabilities, cancers (especially lung cancer, leukemia and lymphoma), sickle cell disease, chronic kidney disease, heart failure, and diabetes appear to be at particularly high mortality risks. (10, 11) Hypertension, obesity, chronic lung diseases, and chronic liver diseases have also been associated with more severe COVID-19 disease. (12-15)

Given their higher mortality risks COVID-19 vaccine allocation policies have prioritized individuals with serious comorbidities. (16) However, since regulatory clinical trials with COVID-19 vaccines excluded those with immunocompromising conditions and those receiving immunosuppressive therapies, few patients with cancer and autoimmune diseases were enrolled. (17, 18) Thus, with limited vaccine safety and efficacy data available, but noting the increased mortality risk, patients with comorbidities may have conflicted COVID-19 vaccine attitudes. We therefore initiated an internet based survey drawing from our health oriented social network to explore issues surrounding COVID-19 vaccine hesitancy in these vulnerable populations. Additionally, we sought to explore early safety experiences among those who had already been vaccinated, as this might provide information useful to combating hesitancy.

## 2. Methods

### 2.1 Study design and participants⍰⍰⍰

Survey participants were⍰identified and⍰recruited from Inspire (www.inspire.com), an⍰online health⍰community⍰of over 2.2⍰million individuals⍰with comorbid conditions and their caregivers. Members anonymously engage with others with similar conditions through discussion posts and direct messaging. The⍰community, with members residing in over 100 countries, represents over 3,600⍰comorbid conditions⍰including cancer, autoimmune diseases, rare diseases and other chronic conditions. As part of enrollment, members are given the opportunity of opting-in to receive research-related surveys directly from Inspire. For this study, email invitations were sent to a computer generated random sample of qualified members on a daily basis. Given the composition of Inspire’s community, survey respondents were not intended to represent a random sampling of the general population or any outside demographic. Prior to taking the survey, all participants completed a consent form that detailed the purpose of the research. Participants were able to⍰withdraw at any time throughout the survey. Duplicate responses were removed by review of unique tokens assigned to participants. Participants were not compensated for completion of the survey. The study protocol and survey were reviewed and approved by the WCG Institutional Review Board (Needham MA) and deemed to be exempt from subject consent.

### 2.2 Measures⍰

The survey consisted of 55-items, with initial responses leading to a potential increase of 8 additional follow-up questions. The survey was⍰implemented using Alchemer, a web-based survey tool. Survey logic, programming, testing, and data validation were done via Alchemer. Items used to assess vaccine perception and⍰hesitancy⍰were adapted from PEW Research Center’s American Trends Panel 2020 survey, with additional questions added and linguistic adjustments. (19) The list of reportable symptoms and effects from the vaccine were adapted from the⍰Pfizer/ BioNTech BNT162b2 mRNA Covid-19 Vaccine FDA Briefing Report. (20) Demographic, health conditions, and treatment related questions were adapted from⍰Inspire’s⍰standard question sets. Behavioral and political view questions were adapted and modified from the Kaiser Family Foundation’s vaccine perception survey.⍰(5)⍰

### 2.3 Statistical Plan⍰

Responses were summarized using descriptive statistics, including percentages and subgroup analysis using crosstabs and Chi Square analysis.⍰Further⍰analysis was conducted using⍰univariate and multivariate⍰General Linear Models.⍰⍰Analysis was conducted using⍰IBM’s⍰SPSS⍰program.

### 2.4 Study Funding

This study was funded by Inspire (Arlington VA) which is responsible for the study design; the collection, analysis, and interpretation of the data; and the decision to approve publication of the finished manuscript.

## 3 Results

### 3.1 Survey Respondent Demographics

Between January 15, 2021 and February 22, 2021, 21,943 individuals completed the questionnaire. The average survey was completed in 17 minutes. The respondent demographic characteristics were similar to the overall Inspire community, although slightly older in age. The median age range of respondents was 56-65 years old with 76% female whereas the Inspire community is 77% female with a median age 40-49 years. Few respondents declared belonging to a racial/ethnic minority. As expected, based on the Inspire community’s target demographics, 100% of participants reported a comorbid condition including 27% with cancer, 23% with autoimmune diseases, and 38% with chronic lung diseases. Respondents were highly educated with 60% holding college or post-graduate degrees. Political leanings were diverse, with 32% self-declaring liberal tendencies, 21% conservative, 24% independent and 23% preferring not to declare. Respondents lived in 123 countries, with 74% residing in the USA, 8% from Canada, 8% from the United Kingdom, 3% from Australia, and the remaining 6% in Europe, Central, South America and the Caribbean, the Middle East, the Russian Federation, Africa and the Far East.

### 3.2 Acceptance of COVID-19 Vaccines

5501 (25%) of the respondents reported that they had already received at least one COVID-19 vaccine injection by February 22, 2021 including 29% of US participants. 688 of the participants from other countries obtained the vaccine including 68% of participants living in Israel, 27% in United Kingdom,4% in Canada, and none in Australia. Additionally, 1462 (7%) had tried but been unable to get it, 9223 (43%) definitely planned to receive a vaccine, and 1029 (5%) indicated that they probably would receive it, leading to an overall vaccine acceptance rate of 81%. (Figure 1)

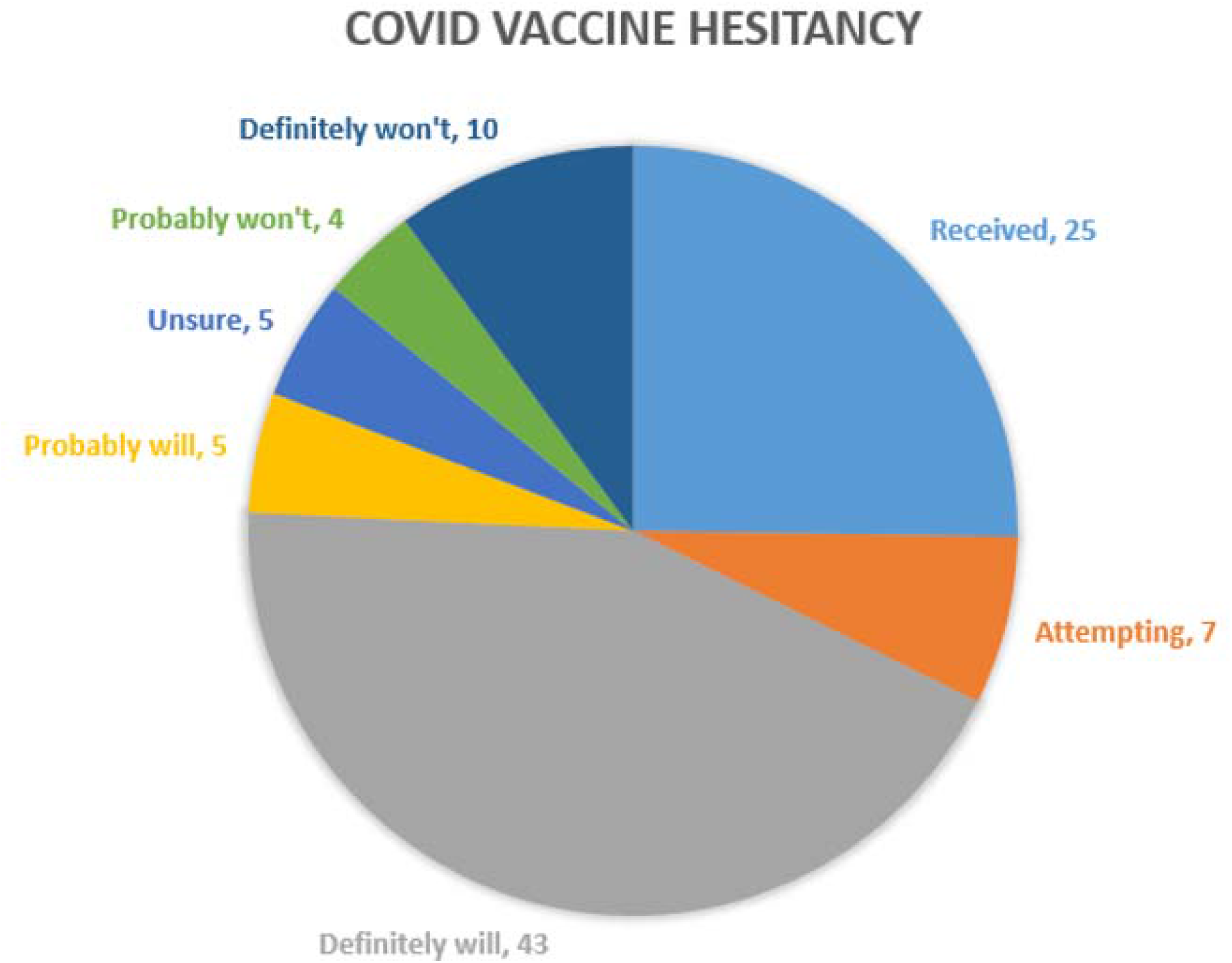

However, 2190 (10%) declared that they would not receive the vaccine, 742 (4%) stated that they probably would not, and 1028 (5%) were not sure whether they would agree to be vaccinated, leading to a vaccine hesitancy rate of 19%. As shown in Table 1, multiple factors by univariate analysis were associated with vaccine hesitancy including younger age, female gender, black-Pacific Island-Native American race/ethnicity, less formal education, and conservative political tendencies. Individuals who do not commonly wear masks, do not receive routine flu vaccinations, and those who felt the vaccines were not developed responsibly or that media coverage has been scientifically inaccurate were more likely to express vaccine hesitancy. Interestingly, individuals who already had contracted COVID-19 (or believed they had unconfirmed infection) were also more likely to decline vaccination.

**Table 1:** Vaccine Hesitancy Rates Among Individuals with Serious Comorbidities.

|  | Vaccine received,<br>Definitely will, or<br>Probably will<br>receive the<br>vaccine | Definitely will<br>not, Probably<br>will not, or<br>unsure about<br>receiving the<br>vaccine | p-value |
| --- | --- | --- | --- |
| **OVERALL** n= 21294; missing 649 | 81% | 19% |  |
| **Age** n=20225 |  |  | **<0.001** |
| <26 n=381 (2%) | 76% | 24% |  |
| 26-35 n=1315 (6%) | 71% | 29% |  |
| 36-45 n=2513 (12%) | 74% | 26% |  |
| 46-55 n=3309 (16%) | 77% | 23% |  |
| 56-65 n=5288 (26%) | 82% | 18% |  |
| 66-75 n=5591 (28%) | 89% | 11% |  |
| 75+ n=1828 (9%) | 90% | 10% |  |
| **Gender** n=20592 |  |  | **<0.001** |
| - male n= 4898 (14%) | 85% | 15% |  |
| - female n= 14,910 (76%) | 82% | 18% |  |
| **Race/ethnicity** n=19465 |  |  | **<0.001** |
| - White n=17354 (85%) | 84% | 16% |  |
| - Black or African American n=514 (2%) | 76% | 24% |  |
| - Hispanic or Latino n=614 (3%) | 83% | 17% |  |
| - Asian n=627 (3%) | 83% | 17% |  |
| -Hawaiian/Pacific Islander n=22 (0.1%) | 68% | 32% |  |
| - Native American/Alaskan n=132 (0.6%) | 67% | 33% |  |
| -Other n=479 (2%) | 70% | 30% |  |
| - Prefer not to answer n=706 (4%) | 43% | 57% |  |
| **Education level** n=17298 |  |  | **<0.001** |
| - high school or less n=1640 (9%) | 76% | 24% |  |
| - vocational and associates degree n=2546 (15%) | 77% | 23% |  |
| - some college n=2914 (17%) | 80% | 21% |  |
| - college degree n=4448 (26%) | 84% | 16% |  |
| - post-graduate n=5927 (34%) | 88% | 12% |  |
| **Political tendency** n=17967 |  |  | **<0.001** |
| - liberal n=5683 (32%) | 95% | 5% |  |
| - conservative n=3711 (21%) | 72% | 24% |  |
| - independent n=4357 (24%) | 81% | 19% |  |
| - prefer not to answer n=4216 (23%) | 76% | 24% |  |
| **Mask policy** n=19468 |  |  | **0.002** |
| -Always/ sometimes wears mask n=18736 (96%) | 82% | 18% |  |
| -Rarely/ Never wears a mask n=732 (4%) | 76% | 24% |  |
| **Routine influenza vaccine** n=20817 |  |  | **<0.001** |
| -Usually gets a flu vaccine n=16269 (78%) | 92% | 8% |  |
| -No flu vaccine n=4548 (22%) | 46% | 54% |  |
| **Media information scientifically accurate**<br>n=19459 |  |  | 0.006 |
| -Yes or generally yes n=10465 (54%) | 82% | 18% |  |
| -No or generally no n=3084(16%) | 79% | 21% |  |
| -Mixed n=5910 (30%) | 82% | 18% |  |
| **Do you trust the vaccine was developed responsibly** answered n=20409 |  |  | **<0.001** |
| -Yes and probably so n=16398 (80%) | 96% | 4% |  |
| -Not sure no=1837 (9%) | 41% | 59% |  |
| -No and probably not n=2174 (11%) | 4% | 96% |  |
| **COVID-19 illness** n=20451 |  |  | **<0.001** |
| Had COVID-19 n=1906 (9%) | 63% | 37% |  |
| Did not have COVID-19 n=17460 (85%) | 84% | 16% |  |
| Unsure if had COVID-19 n=1085 (5%) | 68% | 32% |  |
| **Comorbidities** |  |  |  |
| **Cancer** n=19980 |  |  | **.001** |
| -Yes in treatment n=1463 (7%) | 89% | 11% |  |
| -Yes past treatment n=3996 (20%) | 87% | 13% |  |
| -No Cancer n=14,521 (73%) | 80% | 20% |  |
| **Autoimmune disease n=21294** |  |  | **NS (.078)** |
| -Yes n=4946 (23%) | 80% | 19% |  |
| -No n=16348 (77%) | 82% | 18% |  |
| **Chronic lung disease n=21294** |  |  | **0.03** |
| -Yes n=7544 (35%) | 82% | 18% |  |
| -No n=13750 (65%) | 81% | 19% |  |
| **Smoking status n=18746** |  |  | **0.001** |
| -Current smoker n=993 (5%) | 71% | 29% |  |
| -Past smoker n=6819 (36%) | 23% | 16% |  |
| -Non-smoker n=10934 (58%) | 82% | 18% |  |
| **Hypertension n=21294** |  |  | **NS** |
| -Yes n=5358 (25%) | 82% | 18% |  |
| -No n=15936 (75%) | 81% | 19% |  |
| **Type 2 Diabetes n=21294** |  |  | **NS** |
| -Yes n=1400 (7%) | 81% | 19% |  |
| -No n=19894 (93%) | 81% | 19% |  |
| **Obesity n=21294** |  |  | **NS** |
| - Yes n=3041 (14%) | 80% | 20% |  |
| -No n=18253 (86%) | 82% | 18% |  |

### 3.3 Hesitancy in specific high risk comorbid populations

Among the 5459 individuals with cancer, 703 (11%) of those who were currently receiving treatment were vaccine hesitant as were 538 (13%) of those who had completed prior treatment. Among the 4946 individuals with autoimmune diseases 962 (19%) self-reported vaccine hesitancy as did 1344 (18%) of the 7544 respondents with chronic lung diseases. Similar to cancer patients, individuals with both of these conditions were more likely to accept vaccination. Vaccine hesitancy was also expressed by 20% of those diagnosed as obese, 18% diagnosed with hypertension, and 19% of individuals living with Type 2 diabetes.

### 3.4 Concerns about Vaccines

Of the 3960 respondents who indicated COVID-19 vaccine hesitancy, apprehensions regarding safety of vaccines dominated. 53% indicated a concern that the COVID vaccine is too new, 44% worried about vaccine side effects or discomfort, 33% wanted to see how others responded to the vaccine first, and 8% were concerned about contracting the coronavirus from the vaccine. Issues regarding vaccine development were also common, with 39% citing the role of politics in the development of the vaccine, 9% not trusting the government had insured that the vaccine is safe and effective, and 5% not believing that the vaccine was developed responsibly. Anti-vaccination sentiment was less common, but still noted with 21% not trusting vaccines in general but 40% not trusting the COVID vaccine in particular, and 8% having religious objections to the vaccine. 22% of those with vaccine hesitancy did not believe they needed to be immunized.

### 3.5 Early experience with COVID-19 vaccination in high risk populations

As of the study cutoff, 5501 (25%) survey respondents had received at least one COVID-19 vaccination (Pfizer-BioNTech 48%, Moderna 47%, Oxford-Astra-Zeneca 1%, other/unknown <1%). Following the first injection, 69% had local adverse events (AEs) and 40% self-reported systemic reactions, which increased following the second injection to 77% and 67%, respectively. (Figures 2a, 2b). Of the 5459 cancer patients, 30% had received one injection and 6% completed both vaccine injections. In this cancer population, 65% self-reported local reactions and 34% systemic reactions to the first injection, with local reactions to the second occurring in 72% and 34% systemic reactions. The types of reactions mirrored the overall study population. Of the 5186 individuals with autoimmune disorders, 24% had received one vaccination and 6% had completed the series. In this immunocompromised population local reactions were described in 69% and systemic reactions in 41% with the first, and 78% local and 67% systemic with the second vaccine. Among the 1878 respondents with chronic lung diseases who received the vaccine, 67% self-reported local reactions and 40% systemic reactions to the first injection, with local reactions to the second occurring in 76% and 69% systemic reactions. Similar patterns were noted among respondents with obesity (1^st^ dose 69% local and 43% systemic; 2^nd^ dose 76% local and 75% systemic reactions), hypertension (1^st^ dose 67% local and 39% systemic; 2^nd^ dose 74% local and 66% systemic reactions), and individuals with Type 2 diabetes (1^st^ dose 67% local and 42% systemic; 2^nd^ dose 78% local and 79% systemic reactions).

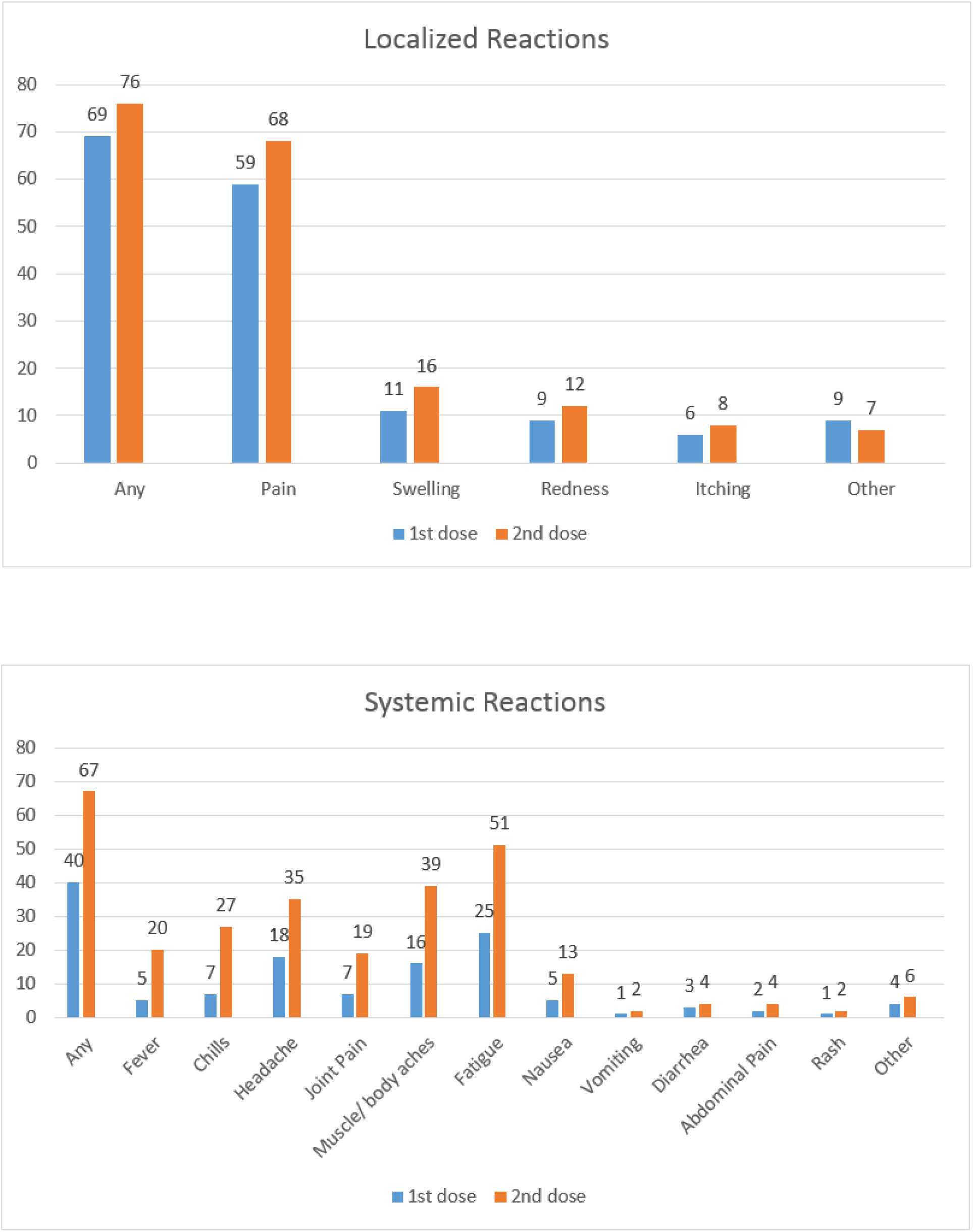

An uncontrolled comparison of the self-reported reaction rates by vaccine type was performed and is detailed in Table 2. A lower rate of systemic reactions during the initial vaccination compared to the clinical trial results, as detailed in the FDA briefing documents, was noted. (20, 21) However, systemic reactions to the booster vaccine in the Inspire population increased dramatically, approaching that of the general trial population.

**Table 2:** Local and Systemic Self-Reported Reactions to COVID-19 Vaccination.

| Self-Reported Reactions | Inspire<br>Pfizer-BioNTech | Inspire<br>Moderna | Regulatory Trial<br>Pfizer-BioNTech<br>(ref 20) | Regulatory Trial<br>Moderna<br>(ref 21) |
| --- | --- | --- | --- | --- |
| 1<sup>st</sup> dose Local | 65% | 75% | 79% | 84% |
| 2<sup>nd</sup> dose Local | 72% | 85% | 73% | 89% |
| 1<sup>st</sup> dose Systemic | 37% | 40% | 59% | 55% |
| 2<sup>nd</sup> dose Systemic | 62% | 77% | 70% | 79% |

## 4 Discussion

In this survey of nearly 22,000 individuals with serious comorbid conditions, eight in ten reported a willingness to receive the COVID-19 vaccine, a rate higher than that reported in public opinion polls drawn from general populations. (4,5) Additionally, 29% of US participants stated that they had already received at least one COVID-19 vaccine injection as of February 22, 2021, which compared favorably to the 18% adult national vaccination rate at that time (p<0.001). (22) These levels of vaccine acceptance appear to confirm a strong desire for protection from SARS-CoV-2 in vulnerable populations, although vaccine allocation prioritization may also be influencing rates.

Unfortunately, almost one in five respondents with comorbid conditions reported COVID-19 vaccine hesitancy. Similar to other studies drawn from the general population, multiple factors were associated with vaccine hesitancy: women, black individuals, residents of rural regions and those with less formal education all expressed vaccine concerns. (5,7) Additionally, respondents who distrusted vaccine development, did not routinely wear masks, had conservative political tendencies and individuals who avoided influenza vaccinations mirrored public opinion polling of vaccine hesitancy. (8, 23) Despite recommendations from the CDC advising vaccination, individuals who had already contracted COVID-19 also avoided vaccination, possibly believing natural immunity alone was protective. (24) Reasons cited for vaccine hesitancy were also similar to general population polling. Since patients with cancer and immune dysfunction were under-represented in vaccine development studies, it might be justified for these groups to have safety concerns. However, distrust of the vaccine development process including speed and politicization of the pandemic were also expressed and are not unique to our comorbid population. (9)

Early experience with vaccinations, as self-reported by the over 5000 respondents who had already been vaccinated, should be reassuring to individuals with serious comorbidities. Side-effect profiles were similar to adverse event reports on the regulatory trials, although overall generally lower in frequency. (20, 21) Whether this is a reflection of the weaker immune status of our population or a result of differences in reporting styles (online survey versus research grade clinical trial monitoring) is unknown. However, an interesting finding was that the prevalence of self-reported systemic reactions to the initial vaccination was much lower than clinical regulatory trials, but increased towards the general population results with the booster, potentially demonstrating an increased value for the second vaccine among immunocompromised patients. Given these adverse event profiles, the recommendations by the CDC to vaccinate individuals with potential immune dysfunction despite a lack of clinical trial data appear justified, although future attention to vaccine efficacy in these populations is required.

## 5 Conclusions

This online survey highlights a high level of acceptance of COVID-19 vaccines among vulnerable individuals with increased mortality risk. However, the finding that still one in five remain vaccine hesitant is of concern and points to a need for additional educational efforts. Similar to surveys drawn from the general population factors associated with hesitancy included younger age, female gender, black-Pacific-Island-Native American heritage, less formal education, conservative political tendencies, resistance to masks or routine influenza vaccinations, and distrust of media coverage. Disinformation about the COVID-19 vaccines is common on social media sites and fosters hesitancy. (25) Our intent is to share our study results with the 2+ million members of the Inspire health community, harnessing the Internet to increase vaccine acceptance by demonstrating vaccine safety among individuals with serious comorbid conditions. A website with daily updated data is available to the general public (www.inspire.com/covid19vax).

## Data Availability

A website, referrenced in the last line of the manuscript, has updated (daily) outcomes of the survey and is available to the public

https://www.inspire.com/covid19vax

## References

1. World Health Organization (WHO). Ten threats to global health in 2019. https://www.who.int/news-room/spotlight/ten-threats-to-global-health-in-2019 (accessed March 1, 2021)

2. Omer S, Salmon DA, Orenstein WA, deHart MP, Halsey N. Vaccine Refusal, Mandatory Immunization, and the Risks of Vaccine-Preventable Diseases. N Engl J Med 2009;360(19):1981

3. World Health Organization (WHO). Coronavirus disease (COVID-19): Herd immunity, lockdowns and COVID-19. https://www.who.int/news-room/q-a-detail/herd-immunity-lockdowns-and-covid-19 (accessed March 1, 2021)

4. Ipsos for the World Economic Forum Davos 21 virtual meeting. Covid-19 vaccines - the world view. https://assets.ipsos-mori.com/worldeconomicforum/Ipsos_vaccine_paper_Jan2021.pdf (accessed March 1, 2021)

5. Kaiser Family Foundation. KFF COVID-19 Vaccine Monitor: February 2021; https://www.kff.org/coronavirus-covid-19/poll-finding/kff-covid-19-vaccine-monitor-february-2021/ (Accessed February 27, 2021)

6. Fisher KA, Bloomstone SJ, Walder L, et al. Attitudes Toward a Potential SARS-CoV-2 Vaccine: A Survey of U.S. Adults. Ann Intern Med. 2020;173:964-973. [Epub ahead of print 4 September 2020]. doi:10.7326/M20-3569

7. Kreps S, Prasad S, Brownstein JS, Hswen Y, Garibaldi BT, Zhang B, Kriner DL. Factors Associated With US Adults’ Likelihood of Accepting COVID-19 Vaccination. JAMA Netw Open. 2020 Oct 1;3(10):e2025594. doi: 10.1001/jamanetworkopen.2020.25594. Erratum in: JAMA Netw Open. 2020 Nov 2;3(11):e2030649. PMID: 33079199; PMCID: PMC7576409.

8. Pogue K, Jensen JL, Stancil CK, et al. Influences on Attitudes Regarding Potential COVID-19 Vaccination in the United States. Vaccines (Basel). 2020;8(4):582. Published 2020 Oct 3. doi:10.3390/vaccines8040582

9. Taylor S, Landry CA, Paluszek MM, Groenewoud R, Rachor GS, Asmundson GJG. A Proactive Approach for Managing COVID-19: The Importance of Understanding the Motivational Roots of Vaccination Hesitancy for SARS-CoV2. Front Psychol. 2020;11:575950. Published 2020 Oct 19. doi:10.3389/fpsyg.2020.575950

10. FAIR Health, West Health Institute, Makary M. Risk Factors for COVID-19 Mortality among Privately Insured Patients. November 11, 2020. https://www.fairhealth.org/publications/whitepapers

11. Dun C, Walsh CM, Bae S, et al. A Machine Learning Study of 534,023 Medicare Beneficiaries with COVID-19: Implications for Personalized Risk Prediction. medRxiv 2020.10.27.20220970; doi: https://doi.org/10.1101/2020.10.27.20220970

12. Liang X, Shi L, Wang Y, et al. The association of hypertension with the severity and mortality of COVID-19 patients: Evidence based on adjusted effect estimates. J Infect. 2020;81(3):e44–e47. doi:10.1016/j.jinf.2020.06.060

13. Tartof SY, Qian L, Hong V, et al. Obesity and Mortality Among Patients Diagnosed With COVID-19: Results From an Integrated Health Care Organization. Ann Intern Med. 2020;173:773-781. [Epub ahead of print 12 August 2020]. doi:10.7326/M20-3742

14. Lippi G, Henry BM. Chronic obstructive pulmonary disease is associated with severe coronavirus disease 2019 (COVID-19). Respir Med. 2020;167:105941. doi:10.1016/j.rmed.2020.105941

15. Kovalic AJ, Satapathy SK, Thuluvath PJ. Prevalence of Chronic Liver Disease in Patients with COVID-19 and Their Clinical Outcomes: A Systematic Review and Meta-Analysis. Hepatology International 14 (July 2020): 612–20, https://doi.org/10.1007/s12072-020-10078-2.

16. Dooling K, Marin M, Wallace M, et al. The Advisory Committee on Immunization Practices’ Updated Interim Recommendation for Allocation of COVID-19 Vaccine — United States, December 2020. MMWR Morb Mortal Wkly Rep 2021;69:1657–1660. DOI: http://dx.doi.org/10.15585/mmwr.mm695152e2externalicon.

17. Polack FP, Thomas SJ, Kitchin N, et al. Safety and Efficacy of the BNT162b2 mRNA Covid-19 Vaccine. N Engl J Med 2020; 383:2603-2615 DOI: 10.1056/NEJMoa2034577

18. Baden LR, El Sahly HM, Essink B, et a. Efficacy and Safety of the mRNA-1273 SARS-CoV-2 Vaccine. N Engl J Med 2021; 384:403-416. DOI: 10.1056/NEJMoa2035389

19. PEW Research Center. 2020 PEW Research Center’s American Trends Panel. https://www.pewresearch.org/science/wp-content/uploads/sites/16/2020/12/PS_2020.12.03_covid19-vaccine-intent_TOPLINE.pdf (accessed March 1, 2021)

20. FDA Briefing Document: Vaccines and Related Biological Products Advisory Committee Meeting, December 10, 2020, Pfizer-BioNTech COVID-19 Vaccine, fda.gov/media/144245/download (accessed February 2, 2021)

21. FDA Briefing Document: Vaccines and Related Biological Products Advisory Committee Meeting, December 17, 2020, Moderna COVID-19 Vaccine, fda.gov/media/144434/download (accessed February 2, 2021)

22. Centers for Disease Control and Prevention. COVID Data Tracker, https://covid.cdc.gov/covid-data-tracker/#vaccinations (accessed February 22, 20121)

23. Ruiz JB, Bell RA. Predictors of intention to vaccinate against COVID-19: Results of a nationwide survey. Vaccine. 2021 Feb 12;39(7):1080–1086. doi: 10.1016/j.vaccine.2021.01.010. Epub 2021 Jan 9. PMID: 33461833; PMCID: PMC7794597.

24. Centers for Disease Control and Prevention. Myths and Facts about COVID-19 Vaccines. https://www.cdc.gov/coronavirus/2019-ncov/vaccines/facts.html (accessed March 1, 2021)

25. Puri N, Coomes EA, Haghbayan H, Gunaratne K. Social media and vaccine hesitancy: new updates for the era of COVID-19 and globalized infectious diseases, Human Vaccines & Immunotherapeutics, 2020; 16:11, 2586–2593, DOI: 10.1080/21645515.2020.1780846

